# Reliability and construct validity of the Technology Device Interference Scale in a sample of children and parents

**DOI:** 10.64898/2026.06.13.26355571

**Authors:** Anett Schumacher, Eric Tu, Katherine T. Cost, Danielle Baribeau, Catherine S. Birken, Alice Charach, Elizabeth Kelley, Christie Burton, Jonathon L. Maguire, Rob Nicolson, Julia Frei, Elisabetta Trinari, Jennifer Crosbie, Daphne J. Korczak

## Abstract

There is increasing interest in parent-child technoference: the interference with personal interactions caused by technology devices. This study examined the reliability and construct validity of the Technology Device Interference Scale (TDIS) to measure technoference in a sample of Canadian parents and children. Parents (n=883) and children (n=376) were recruited from clinical and community settings and completed the TDIS for their own and family member technoference over three timepoints (T1=2023, T2=2024, T3=2025). TDIS internal consistency, test-retest reliability, and construct validity were assessed using Cronbach’s alpha, intraclass correlation coefficient, and confirmatory factor analysis, respectively. The TDIS showed good internal consistency and adequate to good construct validity when used by children to report on their own technoference (all *α*>.70; CFI>.95, TLI>.95, RMSEA<.07) or their parents’ technoference (all *α*>.70; CFI>.95, TLI>.90, RMSEA≤.11). The TDIS had low to acceptable internal consistency and poor model fit for parent report of their own technoference (*α* range: .63 - .66; CFI<.95, TLI≤.80, RMSEA≥.14) or their child’s technoference (*α* range: .56 - .63; CFI<.95, TLI≤.85, RMSEA≥.11); factor loadings for the items “phone” and “tablet” were low (phone<.50, tablet<.50). Test–retest reliability was moderate for all reports. The TDIS demonstrated good reliability and construct validity of child-reported technoference. Parent-reported technoference demonstrated lower reliability and construct validity, with “phone” and “tablet” weakly associated with the other technological devices. Future studies should distinguish between handheld vs. non-handheld devices when investigating parent-reported technoference.

## Introduction

There has been a steady increase in the use of technological devices (cellphones, tablets, laptops) in recent years [1]. Although rates of device use were already on the rise prior to the COVID-19 pandemic, societal lockdowns and school and office closures during the pandemic contributed to the rapid acceleration of daily technological device use by children and their parents over the last five years [2]. In most households, increase in daily technology device usage by children and parents observed during the pandemic has persisted despite pandemic recovery [3,4].

High use of technology devices among children is associated with negative physical and mental health (MH) outcomes [5,6]. Technology device usage at home can also negatively impact the relationship between children and their parents, which may contribute to worsening child MH [7]. While research on parent phubbing (paying attention to mobile phones in place of child interactions) and its negative effects on child MH has increased over the last decade, the impact of overall technology utilization within families is less clear [8,9]. In 2015, McDaniel introduced the term “technoference”, defined as technological device use interruptions of interactions between individuals, to better describe and understand the effects of technological interferences on couple- and parent-child interactions [10]. Emerging evidence suggests that higher technoference in parent-child interactions is associated with increased child screen time, internalizing and externalizing child symptoms, and parent-child relational problems [11–14]. However, studies to date are heterogeneous with respect to included measures, limiting comparability of results.

Two recent scoping reviews highlighted the use of multiple measures to examine parent-child technoference, noting that the Parental Phubbing Scale (PPS), the Generic Scale of Being Phubbed (GSBP), and the Technology Device Interference Scale (TDIS) were the most commonly used tools [7,15]. Both the PPS and GSBP have been validated as reliable tools with good internal consistency and construct validity to measure parental and children’s phubbing [16–18]. However, both scales focus on phubbing, described as distractions in interactions by phone use. The TDIS, on the other hand, measures technoference in relationships across multiple technological devices (i.e., cellphone, television, computer, tablets, iPod, video game console), with higher scores indicating worse technoference [12,19–22]. Berzosa-Grande and team examined the psychometric properties of the TDIS within couple-relationships and confirmed acceptable internal consistency and good construct validity of the scale, but results were limited to technoference in the interactions of adult couples [23]. Studies using the TDIS to investigate technoference in parent-child interactions have adapted the wording of the scale to reflect family interactions, with most studies using parent-report to measure own or child technoference [12,19,20]. Technoference reported by the child has not been well examined; two studies have reported on the perspectives of children with respect to their own and their parents’ technoference [14,21]. To our knowledge, the reliability and construct validity of the TDIS as a measure of technoference from parent and child perspectives has not been investigated previously. However, given the potential importance of technoference in family interactions, a valid and reliable measure of this phenomenon that captures the impacts of multiple different devices, is needed. Thus, the aim of this study was to examine the internal consistency, test-retest reliability, and construct validity of the TDIS in a large sample of children and their parents. We examined parent- and child-reports of each individual’s own and other’s (child and parent, respectively) technoference to allow for comparison of TDIS psychometric properties based on informant.

## Materials and Methods

### Participants and procedures

Data for this study were collected as part of a longitudinal research study of child and adolescent MH in Ontario, Canada, which is comprised of four established cohorts: two clinically referred MH and neurodevelopmental disorder (NDD) cohorts and two community-referred cohorts, followed since April 2020. Further information about the cohorts, consent, and study measures of the larger study have been detailed elsewhere [24]. The current study includes data from three timepoints: Spring 2023 (T1), Spring 2024 (T2), and Spring 2025 (T3). The data were accessed on September 17, 2025, without access to any identifying data. Parents of children ages 3 to 18 years and children ages 10 to 18 years completed annual online surveys via the survey application REDCap [25]. The study was approved by all participating institutional research ethics boards and participants provided informed consent/assent.

### Measures

#### Technoference

Technoference was assessed using the TDIS, which measures the frequency with which technology usage interrupts parent-child interactions [19]. The frequency of technoference across five electronic devices (phone, tablet, television, computer, video game console) was rated by children and parents on a 7-point Likert scale, ranging from 1 (none) to 7 (more than 20 times) per day. TDIS scores were averaged across all five electronic device types, with higher scores indicating more frequent technoference in child-parent interactions [19]. Children ages 10 years and older were asked to report on their own technoference (child-reported child technoference) and on their parent’s technoference (child-reported parent technoference). Parents were asked to report on their own technoference (parent-reported parent technoference) and their child’s technoference (parent-reported child technoference).

#### Demographics and clinical characteristics

Child age, assigned sex, gender identity, ethnicity/ancestry, history of mental MH/NDD diagnosis, and annual household income were collected in November 2022 using items adapted from the Coronavirus Health and Impact Survey questionnaire [26]. Prior MH/NDD diagnosis was defined as one or more of the following diagnoses: anxiety, depression, obsessive–compulsive disorder, learning disorder, attention deficit/hyperactivity disorder, and/or autism spectrum disorder.

### Statistical analysis

Data were cleaned and analyzed using R Statistical Software [27,28]. Repeated measures analysis of variance (RM-ANOVA) was applied to look at change in technoference frequency over time. Pearson’s correlations were conducted to investigate cross-sectional and longitudinal associations between TDIS items for each report (i.e., child-report on child technoference, child-report on parent technoference, parent-report on parent technoference, parent-report on child technoference). To evaluate internal consistency of the TDIS, Cronbach’s alpha (α), corrected item-total correlation (*r*), and Cronbach’s alpha after removing each item (α-i) were calculated for each TDIS report, with α ≥ .70 indicating good reliability and corrected item-total correlation *r* ≥ .30 considered adequate [29,30]. Test–retest reliability of the composite for each of the TDIS was evaluated using Pearson’s correlations and intraclass correlation coefficients (ICCs) across timepoints, with ICC values interpreted using established guidelines (e.g., < .50 = poor, .50–.75 = moderate, .75–.90 = good, > .90 = excellent) [31]. Construct validity of the TDIS, modeled as a one factor model, was assessed using confirmatory factor analysis (CFA) with Diagonally Weighted Least Squares (DWLS) estimator [23,32,33]. Convergent validity was further evaluated using the Average Variance Extracted (AVE), with AVE ≥ .50 indicating that the latent construct explains more variance in its indicators than measurement error [34]. Model fit was assessed with the Comparative Fit Index (CFI ≥ .90 = adequate model fit, CFI ≥ .95 = good model fit), the Tucker- Lewis Index (TLI ≥ .90 = adequate model fit, TLI ≥ .95 = good model fit), the Root Mean Square Error of Approximation (RMSEA ≤ .08 = adequate model fit, RMSEA ≤ .06 = good model fit), and the Standardized Root Mean Squared Residual (SRMR ≤ .09 = adequate model fit, SRMR ≤ .08 = good model fit) [35]. Chi-Square (χ^2^) test results were reported but not used for model fit assessment due to high sensitivity to sample size [36]. Individual factor loadings > .50 were considered acceptable, while factor loadings ≥ .70 were considered strong [37].

## Results

### Participants

The sample consisted of 883 parents and 376 children reporting at T1 (2023). Retention was high over the study timepoints, with 790 (90%) parents and 337 (90%) children participating at T2 (2024), and 780 (88%) parents and 329 (88%) children retained in the study at T3 (2025). The mean age of children with parent-reported outcomes at T1 was 13.3 years (SD = 3.4), the mean age of children with child-report outcomes at T1 was 15.6 years (SD = 2.4). Further details of participant characteristics can be found in Table 1.

**Table 1.** Participant characteristics.

|  | Child report<br>n = 376 | Parent report<br>n = 883 |
| --- | --- | --- |
| **Demographic information at T1** |  |  |
| Age of child - Mean (*SD*) | 15.6 (2.4) | 13.3 (3.4) |
| Assigned sex of child – % (n) |  |  |
| Male | 50.3% (189) | 53.2% (470) |
| Female | 45.2% (170) | 46.7% (412) |
| Did not respond | 4.5% (17) | 0.1% (1) |
| Gender identity<sup>1</sup> of child – % (n) |  |  |
| Cisgender | 83.8% (315) | 95.0% (839) |
| Gender Expansive | 4.5% (17) | 4.5% (40) |
| Did not respond | 11.7% (44) | 0.5% (4) |
| Ethnicity/Ancestry<sup>2</sup> of child – % (n) |  |  |
| European | 61.4% (231) | 67.8% (599) |
| Non-European | 33.0% (124) | 30.8% (272) |
| Did not respond | 5.6% (21) | 1.4% (12) |
| Household income<sup>3</sup> – % (n) |  |  |
| High (≥ 80k) | 54.0% (203) | 59.0% (521) |
| Low ($<$ 80k) | 23.1% (87) | 23.7% (209) |
| Did not respond | 22.9% (86) | 17.3% (153) |
| Prior MH/NDD diagnosis<sup>4</sup> of child – % (n) |  |  |
| Yes | 40.4% (152) | 36.2% (320) |
| No | 44.4% (167) | 54.5% (481) |
| Did not respond | 15.2% (57) | 9.3% (82) |
| **Frequency of technoference<sup>5</sup> across all time points (T1 – T3)** |  |  |
| Child technoference - Mean (*SD*) | **Child report** | **Parent report** |
| T1 | 2.05 (1.05) | 2.10 (0.88) |
| T2 | 1.93 (0.95) | 2.09 (0.91) |
| T3 | 1.92 (0.97) | 2.05 (0.86) |
| Parent technoference - Mean (*SD*) | **Child report** | **Parent report** |
| T1 | 2.01 (0.90) | 1.88 (0.70) |
| T2 | 1.89 (0.86) | 1.85 (0.68) |
| T3 | 1.94 (0.93) | 1.81 (0.70) |

Note. <sup>1</sup>Gender: Cisgender = Cisgender Male, Cisgender Female; Gender Expansive = Transgender Boy, Transgender Girl, Non-Binary, Gender-Fluid, Two-Spirit. <sup>2</sup>Ethnicity/Ancestry: Non-European = Indigenous, Caribbean, African, Asian, Oceanian, South-American, Other. <sup>3</sup>Household Income: High Annual Household Income = ≥ \$80,000 CAD. <sup>4</sup>Prior MH/NDD diagnosis = prior mental health/neurodevelopmental disorder history, including anxiety, depression, obsessive–compulsive disorder, learning disorder, attention deficit/hyperactivity disorder, and/or autism spectrum disorder. <sup>5</sup>Technoference frequency was measured using the Technology Device Interference Scale (TDIS) and rated on a 7-point Likert scale, ranging from 1 (none) to 7 (more than 20 times) per day.

### Descriptives of the TDIS

Child and parent technoference for all three time points is presented in Table 1. Four separate omnibus RM-ANOVA tests did not indicate a statistically significant difference in child or parent technoference across time points (child-reported child technoference F(587) = 3.09, *p* = .05); parent-reported child technoference: F(1468) = 1.20, *p* = .30; parent-reported parent technoference: F(1456) = 2.21, *p* = .11; child-reported parent technoference F(604) = 2.24, *p* = .11). No post hoc comparison between timepoints was conducted. Cross-sectional and longitudinal correlations between TDIS items are shown in S1-S4 Tables. Cross-sectionally, all TDIS items were significantly correlated with each other for child technoference when reported by child, and for parent technoference when reported by child or parent (all *r*’s between .09 - .55). However, the TDIS items “phone” and “tablet” were not correlated for child technoference at T1 when reported by parent (*r* = .02, S2 Table). Longitudinally, each TDIS item was correlated between time points, irrespective of informant or who was being reported on. Test–retest reliability analyses indicated moderate temporal stability of the TDIS composite scores for all types (i.e., correlations between time points ranged from .39 to .50, and ICCs across time points ranged from .44 to .48; see S5 Table).

### Internal consistency of the TDIS

Internal consistency, including Cronbach’s alphas (α), corrected item-total correlations (*r*), and Cronbach’s alpha after removing each item (α-i), for all four TDIS reports within each time point are reported in Table 2.

**Table 2.** Internal consistency indices for items of the Technology Device Interference Scale at T1, T2, and T3.

| Item | Child-reported<br>Child technofence |  | Child-reported<br>Parent technofence |  | Parent-reported<br>Parent technofence |  | Parent-reported<br>Child technofence |  |
| --- | --- | --- | --- | --- | --- | --- | --- | --- |
| T1 | $r$ | $\alpha - i$ | $r$ | $\alpha - i$ | $r$ | $\alpha - i$ | $r$ | $\alpha - i$ |
| Total Technoference | $\alpha = 0.76$,<br>*CI* [.73, .78] | | $\alpha = 0.72$,<br>*CI* [.69, .75] | | $\alpha = 0.63$,<br>*CI* [.60, .67] | | $\alpha = 0.56$,<br>*CI* [.52, .61] | |
| Phone | 0.53 | 0.74 | 0.58 | 0.67 | 0.45 | 0.60 | 0.37 | 0.56 |
| Tablet | 0.60 | 0.72 | 0.54 | 0.68 | 0.49 | 0.60 | 0.31 | 0.56 |
| Television | 0.71 | 0.69 | 0.63 | 0.65 | 0.53 | 0.56 | 0.47 | 0.51 |
| Computer | 0.69 | 0.68 | 0.69 | 0.62 | 0.55 | 0.54 | 0.62 | 0.40 |
| Video game console | 0.53 | 0.73 | 0.39 | 0.72 | 0.53 | 0.60 | 0.52 | 0.48 |
| **T2** | ***r*** | **$\alpha - i$** | ***r*** | **$\alpha - i$** | ***r*** | **$\alpha - i$** | ***r*** | **$\alpha - i$** |
| Total Technoference | $\alpha = 0.75$,<br>*CI* [.73, .78] | | $\alpha = 0.76$,<br>*CI* [.74, .78] | | $\alpha = 0.65$,<br>*CI* [.61, .68] | | $\alpha = 0.64$,<br>*CI* [.60, .67] | |
| Phone | 0.50 | 0.75 | 0.63 | 0.71 | 0.42 | 0.63 | 0.36 | 0.65 |
| Tablet | 0.58 | 0.73 | 0.64 | 0.72 | 0.48 | 0.61 | 0.41 | 0.61 |
| Television | 0.65 | 0.70 | 0.62 | 0.71 | 0.55 | 0.56 | 0.57 | 0.55 |
| Computer | 0.72 | 0.67 | 0.65 | 0.70 | 0.57 | 0.54 | 0.64 | 0.50 |
| Video game console | 0.62 | 0.71 | 0.55 | 0.75 | 0.57 | 0.59 | 0.55 | 0.55 |
| **T3** | ***r*** | **$\alpha - i$** | ***r*** | **$\alpha - i$** | ***r*** | **$\alpha - i$** | ***r*** | **$\alpha - i$** |
| Total Technoference | $\alpha = 0.75$,<br>*CI* [.73, .78] | | $\alpha = 0.74$,<br>*CI* [.72, .77] | | $\alpha = 0.68$,<br>*CI* [.65, .71] | | $\alpha = 0.61$,<br>*CI* [.57, .65] | |
| Phone | 0.57 | 0.77 | 0.60 | 0.72 | 0.44 | 0.65 | 0.33 | 0.62 |
| Tablet | 0.55 | 0.77 | 0.58 | 0.73 | 0.53 | 0.62 | 0.41 | 0.58 |
| Television | 0.68 | 0.73 | 0.59 | 0.72 | 0.64 | 0.56 | 0.55 | 0.53 |
| Computer | 0.73 | 0.71 | 0.73 | 0.67 | 0.54 | 0.59 | 0.62 | 0.47 |
| Video game console | 0.66 | 0.74 | 0.59 | 0.73 | 0.52 | 0.63 | 0.52 | 0.53 |

Note. $\alpha$ = Cronbach's alpha ( $\alpha \geq .70$ = good reliability), 95% Confidence Interval calculated using the [43] method; $\alpha - i$ = Cronbach's alpha if item is removed; $r$ = corrected item-total correlation ( $r \geq .30$ = adequate reliability).

#### TDIS internal consistency for child-reports

Internal consistency for the TDIS was good within each time point when children reported on child and parent technoference (all α > .70), with adequate corrected item-total correlations for all items (all *r* > .30).

#### TDIS internal consistency for parent-reports

Internal consistency for the TDIS was acceptable within each time point when parents reported on parent technoference (all α > .60), and corrected item-total correlations was adequate for all items (all *r* > .30). Removal of individual items did not lead to an increase in scale reliability. Internal consistency for the TDIS when parents reported on child technoference was low at T1 (α = .56) and increased over time (T2: α = .63, T3: α = .60). Removal of the item “phone” led to an increase in scale reliability at T2 (α-i = .65) and T3 (α-i = .62) but not at T1 (α-i = .56). Corrected item-total correlations for all items were adequate (all *r* > .30) over time.

### Construct validity of the TDIS

Fit indices for all models are shown in Table 3, while individual factor loadings for all TDIS reports within each time point are reported in Table 4.

**Table 3.** Fit indices for confirmatory factor analyses models of the Technology Device Interference Scale at T1, T2, and T3.

| Fit Index | Child-reported<br>Child technoference | Child-reported<br>Parent technoference | Parent-reported<br>Parent technoference | Parent-reported<br>Child technoference |
| --- | --- | --- | --- | --- |
| **T1** |  |  |  |  |
| CFI | 0.987 | 0.957 | 0.822 | 0.800 |
| TLI | 0.974 | 0.914 | 0.645 | 0.600 |
| RMSEA | 0.068 | 0.114 | 0.173 | 0.167 |
| SRMR | 0.036 | 0.058 | 0.101 | 0.090 |
| $\chi^2$ (df) | 13.7(5); $p = .018$ | 29.2(5); $p < .001$ | 135.9(5); $p < .001$ | 128.7(5); $p < .001$ |
| **T2** |  |  |  |  |
| CFI | 0.994 | 0.971 | 0.888 | 0.925 |
| TLI | 0.988 | 0.942 | 0.777 | 0.850 |
| RMSEA | 0.050 | 0.109 | 0.140 | 0.118 |
| SRMR | 0.028 | 0.057 | 0.083 | 0.060 |
| $\chi^2$ | 9.9(5); p = .079 | 28.4(5); p < .001 | 90.9(5); p < .001 | 66.0(5); p < .001 |
| **T3** |  |  |  |  |
| CFI | 0.996 | 0.983 | 0.904 | 0.885 |
| TLI | 0.993 | 0.967 | 0.808 | 0.771 |
| RMSEA | 0.040 | 0.084 | 0.140 | 0.135 |
| SRMR | 0.027 | 0.048 | 0.088 | 0.070 |
| $\chi^2$ | 8.0(5); p = .155 | 18.0(5); p = .003 | 89.1(5); p < .001 | 83.3(5); p < .001 |

*Note.* Scaled estimates are reported. CFI = Comparative Fit Index (CFI ≥ .90 = adequate model fit, CFI ≥ .95 = good model fit); TLI = Tucker- Lewis Index (TLI ≥ .90 = adequate model fit, TLI ≥ .95 = good model fit); RMSEA = Root Mean Square Error of Approximation (RMSEA ≤ .08 = adequate model fit, RMSEA ≤ .06 = good model fit); SRMR = Standardized Root Mean Squared Residual (SRMR ≤ .09 = adequate model fit, SRMR ≤ .08 = good model fit); $\chi^2$ = Chi-Square test.

**Table 4.** Individual factor loadings for items of the Technology Device Interference Scale at T1, T2, and T3.

| Item | Child-reported<br>Child technofence | Child-reported<br>Parent technofence | Parent-reported<br>Parent technofence | Parent-reported<br>Child technofence |
| --- | --- | --- | --- | --- |
| **T1** |  |  |  |  |
| Phone | 0.57 | 0.60 | 0.40 | 0.40 |
| Tablet | 0.66 | 0.59 | 0.60 | 0.32 |
| Television | 0.76 | 0.68 | 0.61 | 0.55 |
| Computer | 0.75 | 0.79 | 0.61 | 0.69 |
| Video<br>game<br>console | 0.63 | 0.51 | 0.73 | 0.60 |
| **T2** |  |  |  |  |
| Phone | 0.58 | 0.72 | 0.46 | 0.35 |
| Tablet | 0.71 | 0.72 | 0.59 | 0.45 |
| Television | 0.71 | 0.71 | 0.61 | 0.67 |
| Computer | 0.79 | 0.70 | 0.66 | 0.69 |
| Video game console | 0.73 | 0.69 | 0.77 | 0.66 |
| **T3** |  |  |  |  |
| Phone | 0.61 | 0.69 | 0.47 | 0.31 |
| Tablet | 0.59 | 0.65 | 0.63 | 0.45 |
| Television | 0.74 | 0.66 | 0.71 | 0.65 |
| Computer | 0.81 | 0.82 | 0.63 | 0.69 |
| Video game console | 0.74 | 0.76 | 0.78 | 0.64 |

*Note.* Individual factor loadings $> .50$ were considered acceptable, individual factor loadings $\geq .70$ were considered strong.

#### TDIS construct validity for child-reports

The TDIS used by children to report on child technoference showed good model fit and acceptable to strong item factor loadings (range: .57 - .81) and moderate AVE (range: .46 - .50) within each time point. The TDIS used by children to report on parent technoference showed adequate model fit at T1 and T2 and good model fit at T3, while AVE was moderate for all time points (range: .46 - .50). Item factor loadings were acceptable to strong within each time point (range: .51 - .82).

#### TDIS construct validity for parent-reports

Parent-report of parent technoference using the TDIS showed poor model fit and poor AVE (range: .36 - .43) within each time point. Most item factor loadings were acceptable to strong within each time point (range: .59 - .78) except for the item “phone” (< .50). Parent-report of child technoference using the TDIS had similarly poor model fit and poor AVE (range: .28 - .34) within each time point. Factor loadings for items “television”, “computer”, and “video game console” were acceptable within each time point (range: .55 - .69), while factor loadings for the items “phone” and “tablet” were poor at T1, T2 and T3 (phone ≤ .40, tablet < .50).

## Discussion

This study examined the reliability and construct validity of the TDIS to measure parent-child technoference in a sample of Canadian children and their parents. Using both child and parent perspectives on own vs. family member technoference allowed for investigation of potential differences in the use of the TDIS across family members. Frequency of technoference did not change across timepoints, irrespective of informant, indicating low variability of technoference over time. Child-report of technoference using the TDIS showed good internal consistency, good model fit, and adequate factor loadings within each of the three timepoints when reporting on their own or their parents’ technoference. In contrast, parent-report of technoference using the TDIS showed low to acceptable internal consistency, poor model fit, and poor factor loadings for the item “phone” and “tablet” within each timepoints when reporting on their own or their child’s technoference. Test-retest reliability showed moderate stability for all TDIS reports.

Internal consistency and construct validity of the TDIS was good when children used it to report on technoference, irrespective of type of technoference (child vs. parent), suggesting that the scale was able to measure technoference when children reported on it. Past research used adapted, non-validated versions of the TDIS to examine child-reported technoference [14,21]. To our knowledge, this is the first study to demonstrate that the TDIS is a reliable and valid measure to examine technoference when reported by children. Furthermore, the TDIS showed moderate test-retest reliability for both child reports, indicating that it is a reliable measure of both own and parent technoference.

In contrast, parent-report using the TDIS showed weaker internal consistency and construct validity when parents assessed their own and their child’s technoference, suggesting that parent-report TDIS values may require caution in interpretation. The phone item had consistently poor factor loadings across time, indicating a weak correlation between this item and the technoference construct. However, removal of the phone item did not improve internal consistency of the scale. The TDIS used by parents to report on their child’s technoference had lower internal reliability at the first timepoint compared to the following timepoints. Across all timepoints, model fit was poor, and factors loadings for the phone and tablet items were weak. The removal of the phone item slightly improved reliability at later timepoints. The results of this study indicate that the TDIS may not reliably measure technoference across devices when reported by parents. Previous research has shown that social desirability bias can be an important contributor to parental responses in research studies [38]. It is thus possible that parents in this study may have been more susceptible to responding in a socially desirable way to the TDIS items about themselves and their children. Alternatively, parents in general may be less observant or accurate in identifying technoference, leading to lower model fit. Future studies are needed to examine parents’ thoughts and knowledge of technoference to better understand if these may bias the way parents report on technoference.

TDIS items referring to parent phone and tablet use had low factor loadings, indicating that these items were weakly associated with those assessing technoference as a result of other device use. Previous research has shown that the use of phones and tablets (handheld devices) has increased over the last years, compared with the stable use of television and decreased use of computer and video game console [39–41]. Furthermore, a recent study investigating the effects of parent-reported usage of handheld devices vs. television found an association between high parental usage of handheld devices and an increase in child screen time [40]. Thus, parents may consider phone and tablet use differently than they do other electronic devices, leading to weak correlations between TDIS rated handheld vs. non-handheld devices. In detail, it is possible that parents report more frequent own- and child technoference due to phones and tablets as a result of increased use relative to other devices. Thus, examining TDIS scores separately by device for parent-reports may be more informative in the assessment of technology interruptions in families compared with use of the overall score, and should be considered in future research.

### Strengths and Limitations

This study has several strengths. First, reliability and construct validity of the TDIS was explored using a robust sample size of children and their parents (total of 1259 participants). Second, three consecutive timepoints were used to examine parent- and child-reports, indicating moderate test-retest properties of the TDIS. However, there are also limitations to consider. This study focussed mainly on children and parents with higher socioeconomic backgrounds, limiting the results to families with disproportionally greater economic resources. Further research across different socioeconomic backgrounds is needed to examine if the reliability and construct validity of the TDIS is depended on socioeconomic conditions. Second, child-reports were limited to older children, ages 10 years and up. In general, children 8 years and older are able to complete age-appropriate questionnaires [42]. Past research investigating child-reported technoference used TDIS child-reports from children 10 years and older and found associations between technoference and mental health [14,21]. Thus, future studies are encouraged to confirm the reliability and construct validity of the TDIS when using reports from children younger than 10 years.

## Conclusion

The TDIS showed good reliability and construct validity when reported by children on their own and their parents’ technoference. However, psychometric properties were weaker when reported by parents, indicating that parent and children may perceive technoference differently. The handheld devices “phone” and “tablet” were weakly associated with non-handheld technological devices when parents reported on technoference. Thus, future research is encouraged to distinguish between the usage of handheld vs. non-handheld devices when investigating parent-reported technoference.

## Data Availability

The data that support the findings of this study are not openly available as they include sensitive and confidential information. Data are available from the corresponding author upon reasonable request (including a study outline), subject to review.

## Acknowledgements

The authors acknowledge the families, children, and youth who have generously contributed their time and their experience participating in this research. The authors also acknowledge the trainees, analysts, project coordinators, and cohort staff whose dedication has made this research possible.

## Supporting information

**S1 Table. Cross-sectional and longitudinal correlations between TDIS items for child-reported child technoference.** *Note*. M and SD are used to represent mean and standard deviation, respectively. * indicates p < .05. ** indicates p < .01.

**S2 Table. Cross-sectional and longitudinal correlations between TDIS items for parent-reported child technoference.** *Note*. M and SD are used to represent mean and standard deviation, respectively. * indicates p < .05. ** indicates p < .01.

**S3 Table. Cross-sectional and longitudinal correlations between TDIS items for parent-reported parent technoference.** *Note*. M and SD are used to represent mean and standard deviation, respectively. * indicates p < .05. ** indicates p < .01.

**S4 Table. Cross-sectional and longitudinal correlations between TDIS items for child-reported parent technoference.** *Note*. M and SD are used to represent mean and standard deviation, respectively. * indicates p < .05. ** indicates p < .01.

**S5 Table. Test-retest reliability of the TDIS composite for different technoference reports across timepoints.** *Note*. Intraclass Correlation (ICC) was assessed using single-measure, absolute agreement. *** indicates p < .001.

